# HeadSOAR, a Multicenter, Randomized, Open-Label Trial of Head Positioning After Endovascular Therapy for Large Vessel Occlusion Stroke: Study Rationale and Design

**DOI:** 10.1101/2025.10.30.25339117

**Authors:** Zhengzhou Yuan, Jianhua Peng, Long Gu, Gaoming Li, Thanh N. Nguyen, Changwei Guo, Jinglun Li, Zhenghua Zhou, Chong Zheng, GuoYong Zeng, Bin Wang, Weizheng Xie, Xiangping Cheng, Haoyue Wang, Bin Han, Tao Hao, Jian Wang, Duolao Wang, Kecheng Guo, Tianqi Tu, Xingxia Wang, Xiu Chen, Yuan Yang, Renliang Meng, Shuang Qiu, Li Jiang, Wenjie Zi, Yong Jiang, the HeadSOAR Investigators

**Author notes:** Correspondence to: Yong Jiang, MD. Department of Neurosurgery, the Affiliated Hospital, Southwest Medical University, No. 25 Taiping Street, Jiangyang District, Luzhou, 646000, China., Wenjie Zi, MD. Department of Neurosurgery, Xinqiao Hospital and The Second Affiliated Hospital, Army Medical University (Third Military Medical University), Chongqing, 400037, China. Zhengzhou Yuan, Jianhua Peng, Long Gu, Gaoming Li are co-first authors. Both Yong Jiang and Wenjie Zi are co-senior authors.

## Abstract

**Background:** The elevated head position may lower intracranial pressure but has been associated with an increased risk of early neurological deterioration before endovascular therapy (EVT). However, among patients with medium-to-large infarct cores who achieve successful reperfusion after EVT, the efficacy and safety of elevated head positioning remain uncertain.

**Objective:** This study aims to evaluate whether maintaining patients in an elevated head position (30°-40°) versus a flat lying position (0°-10°) for 72 hours after EVT improves functional outcomes at 90 days.

**Methods and design:** The Head Positioning After Endovascular Therapy for Acute Ischemic Stroke (HeadSOAR) trial is an investigator-initiated, multicenter, prospective, randomized, open-label, blinded-end point (PROBE) trial. A total of 1,332 patients with acute anterior circulation LVO, medium to large core strokes, successful reperfusion (eTICI ≥2b) after EVT, will be randomized in a 1:1 ratio to either an elevated head position (30°-40°) or flat lying position (0°-10°) for 72 hours.

**Outcomes:** The primary outcome is the distribution of modified Rankin Scale (mRS) scores at 90 days. Secondary outcomes include the proportions of patients with mRS scores of 0–1, 0–2, and 0–3, as well as the European Quality Visual Analogue Scale (EQ-VAS) score at 90 days. Safety endpoints include 90-day mortality and adverse events. The study will be conducted in more than 60 centers in China, with a planned completion in 2025.

**Conclusion:** The HeadSOAR trial will provide evidence on whether elevated head positioning after EVT influences functional recovery.

**Trial registry number:** NCT06115707 (www.clinicaltrials.gov).

## INTRODUCTION

Stroke is one of the leading causes of disability and mortality worldwide, particularly in patients with large vessel occlusion (LVO) ^1^. Endovascular therapy (EVT) has emerged as the standard treatment for LVO, achieving successful reperfusion rates exceeding 80%^2^. However, poor outcomes persist in 54-70% of patients who receive EVT, with mortality rates ranging from 15% to 35% within 90 days post-EVT^3^. Complications such as pulmonary infections or cerebral edema account for a substantial proportion of these deaths^4^, highlighting the importance of strategies to mitigate these risks and improve outcomes.

Head positioning after acute stroke is a potentially impactful yet underexplored intervention. Guidelines and expert consensus advocate for elevating the head to prevent pulmonary infections and cerebral edema by decreasing aspiration risk and reducing intracranial pressure^5,6^. However, the optimal head position remains uncertain^7^. Although lying flat has been associated with increased cerebral blood flow to ischemic areas, this has not translated to improved clinical outcomes, as evidenced by the HeadPoST trials and related meta-analyses^8,9^. A phase 2 randomized trial of Trendelenberg position in patients with acute moderate ischemic stroke related to large artery atherosclerosis showed numerically higher rates of favorable outcome (65% vs. 50%) at 90 days^10^. Conversely, elevated head positions may benefit specific subgroups of patients, such as patients with malignant middle cerebral artery infarction or primary intracerebral hemorrhage (ICH)^11^. EVT-treated patients represent a unique subgroup of patients with more severe strokes, longer hospital stays, and a higher risk of pulmonary infections, affecting up to half of patients^12^. Additionally, cerebral edema occurs in approximately 40% of these patients^13^. These challenges underscore the need for targeted post-EVT management strategies.

We designed the Randomized Trial of Head Positioning After Endovascular Therapy for Acute Ischemic Stroke (HeadSOAR) to investigate whether adopting head elevation after EVT could reduce the risk of complications and improve 90-day functional outcomes compared with the flat lying position.

## METHODS

### Design

The HeadSOAR trial is an investigator-initiated, multicenter, prospective, randomized, open-label trial with blinded evaluation of outcomes (PROBE design) clinical trial, aiming to investigate the efficacy and safety of head elevation on outcomes following

EVT for acute ischemic stroke. The trial is registered at www.clinicaltrials.gov (NCT06115707). The study protocol adheres to the SPIRIT (Standard Protocol Items: Recommendations for Interventional Trials) statement^14^ and complies with the Declaration of Helsinki.^15^ The trial has been approved by the Ethics Committee of the Southwest Medical University and all participating centers. The trial will be conducted at more than 60 hospitals in China with prior experience conducting acute ischemic stroke (AIS) trials with the capacity to perform both intravenous thrombolysis (IVT) and EVT, and with more than 30 cases of EVT for AIS annually. All participating neurointerventionalists must have more than 3 years of experience in neurointervention and have performed at least 30 EVT procedures. The study recruited its first patient in November 2023 and is expected to end in May 2025. The flowchart of HeadSOAR trial is shown in Figure 1

**Figure 1.**
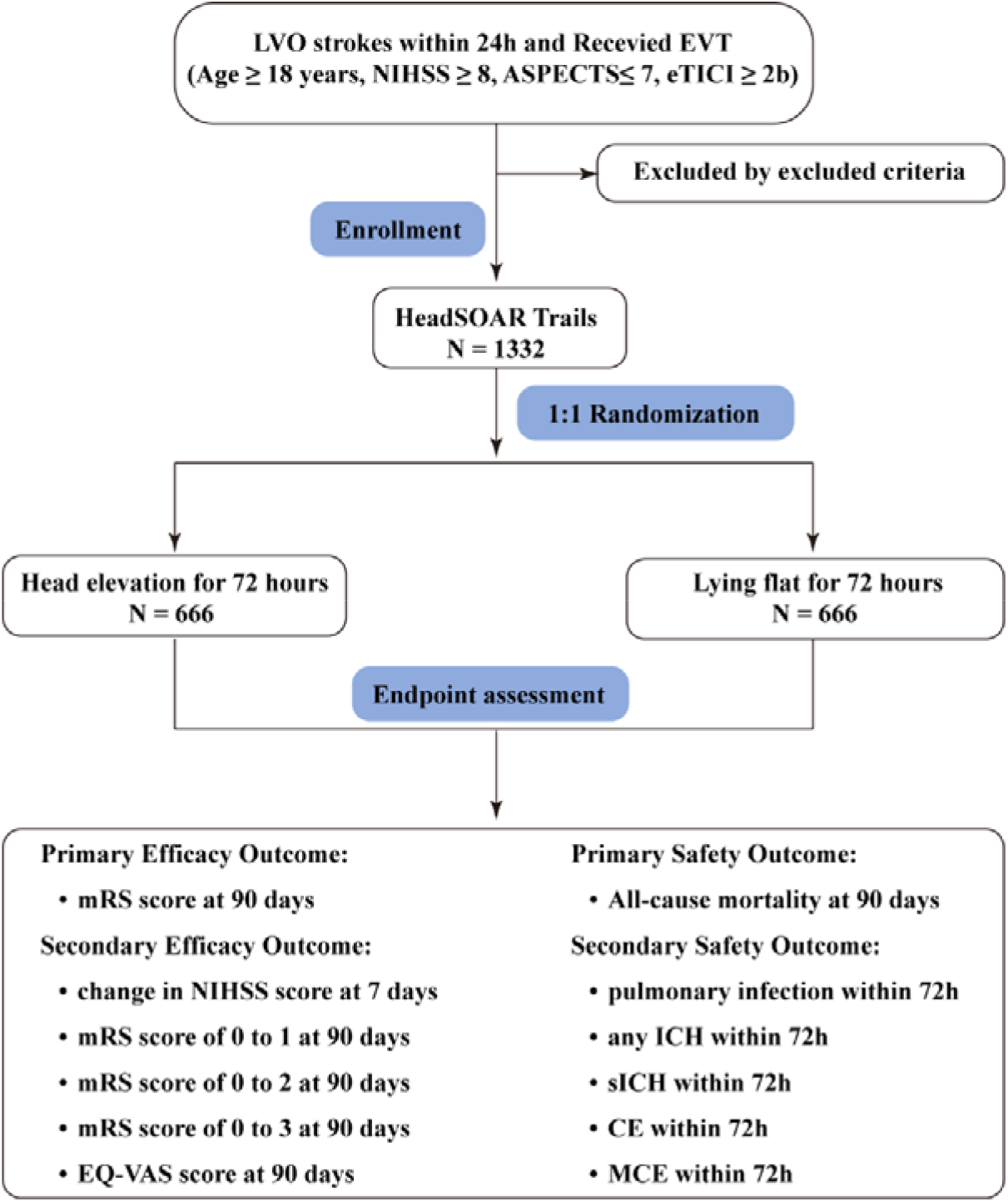
Study flowchart. LVO, large vessel occlusion; EVT, endovascular therapy; NIHSS, National Institute of Health Stroke Scale; ASPECTS, Alberta Stroke Program Early Computed Tomography score; eTICI, expanded Thrombolysis in Cerebral Infarction; ICH, intracranial cerebral hemorrhage; sICH, symptomatic intracranial hemorrhage; mRS, modified Rankin scale; CE, cerebral edema; MCE, malignant cerebral edema; EQ-VAS, European Quality Visual Analogue Scale.

The study inclusion criteria are as follows:

1. clinical signs consistent with acute ischemic stroke;
2. Age ≥18 years old;
3. Proved anterior circulation large vessel (ICA, M1, M2) occlusion on digital subtraction angiography with/without cervical lesion (tandem);
4. NIHSS score ≥ 8 points before endovascular treatment;
5. ASPECTS score ≤ 7 points before endovascular treatment;
6. Successful vessel recanalization after endovascular treatment (defined as an eTICI score of 2b, 2c, or 3);
7. The time from onset to randomization ≤ 24 hours (the onset time is defined as the last normal time);
8. Written informed consent is obtained from patients and/or their legal representatives.

The study exclusion criteria are:

1. pre-stroke mRS score>1 point;
2. Patients with acute occlusions in multiple vascular territories (e.g., bilateral anterior circulation middle cerebral artery, or anterior/posterior circulation);
3. Currently pregnant or lactating or serum beta HCG test is positive on admission;
4. Contraindications to a flat head position;
5. Any terminal illness with life expectancy less than 6 months;
6. Participating in other clinical trials;
7. Patients with a preexisting neurological or psychiatric disease that would confound the neurological functional evaluations;
8. Unlikely to be available for 90-day follow-up.

### Randomization

Following confirmation of eligibility and the provision of informed consent, randomization will be conducted through a web-based application WeChat Mini Program (https://mp.weixin.qq.com/). Permuted block randomization stratified by participating center will be used to generate the randomization list. Eligible patients will be randomly assigned to either the lying flat (0°-10°) or head elevation (30°-40°) head position group in a 1:1 ratio. Randomization will be conducted after EVT as soon as possible.

### Blinding

As HeadSOAR is an open label blind endpoint trial, treatment allocation is known to the treating physicians and the patients. The imaging and endpoint assessments conducted during the trial are carried out by investigators who are independent of the trial allocation, in accordance with the standardized recording form and assessment process designed for this study. The statistician who performs the statistical analyses of the data will be blinded to the intervention. The members of the Steering Committee are blinded regarding the efficacy and safety analyses.

### Intervention

EVT procedures in the HeadSOAR study will be based on the 2019 AHA/ASA Guidelines for the Early Management of Acute Ischemic Stroke^7^ and the Chinese Guidelines for Endovascular Treatment of Acute Ischemic Stroke (2023)^16^. The specific treatment modality of EVT for each patient will be at the discretion of the treatment team, which may include stent retriever thrombectomy, aspiration thrombectomy, balloon angioplasty, stent placement, or a combination of these methods.

Patients eligible for the trial will be randomly grouped into either the head elevation (30°-40°) or lying flat (0°-10°) head position group as soon as possible after randomization and will be maintained in their assigned positions for 72 hours. Patients in both groups will be permitted four periods of time each day to change position at their discretion, for activities such as eating, undergoing tests, and getting out of bed for rehabilitation. Each period will not exceed one hour. Adherence to the assigned position will be monitored through direct observation by caregivers, nurses, and other healthcare providers, who are informed of the patient’s allocation. In addition to the differences in head-up interventions, standard pharmacological treatment and nursing care after acute stroke will be employed in both groups according to the Chinese Guidelines for Endovascular Treatment of Acute Ischemic Stroke (2023)^16^.

### Study schedule

Data collection will include baseline characteristics, medical history, laboratory findings, stroke severity (measured by NIHSS), pretreatment and posttreatment imaging findings, EVT characteristics, complications, and functional outcomes at 90 days. Key time metrics such as onset-to-treatment time will also be recorded. Table 1 provides a brief assessment flowchart.

**Table 1.** Schedule of Assessments.

|  | Visit (Time in hours / days) |  |  |  |  |  |
| --- | --- | --- | --- | --- | --- | --- |
|  | Screening |  | Treatment | Follow Up |  |  |
|  | V0 | R | V1 <sup>a</sup> | V2<br>(72h±12h) | V3 <sup>b</sup><br>(5-7d) | V4<br>(90±7d) |
| Informed consent | X |  |  |  |  |  |
| Demographic data | X |  |  |  |  |  |
| Medical history | X |  |  |  |  |  |
| Physical examination | X |  |  | X | X |  |
| Pre-stroke mRS | X |  |  |  |  |  |
| NIHSS | X |  |  | X | X |  |
| Previous medication | X |  |  |  |  |  |
| Blood pressure and heart rate | X |  | X<sup>c</sup> |  |  |  |
| Local laboratory result | X |  |  | X |  |  |
| Pregnancy test <sup>d</sup> | X |  |  |  |  |  |
| Brain CT/MRI plus CTA/MRA/DSA | X |  |  |  |  |  |
| Chest X-ray/CT scan | X |  |  | X<sup>e</sup> |  |  |
| Brain CT/MRI scan |  |  |  | X<sup>f</sup> |  |  |
| Inclusion/exclusion criteria | X | X |  |  |  |  |
| Randomization |  | X |  |  |  |  |
| Head position intervention <sup>g</sup> |  |  | X | X |  |  |
| Concomitant medication | X |  | X | X | X | X |
| 12-lead ECG | X |  |  |  |  |  |
| Adverse events |  |  | X | X | X | X |
| mRS | X |  |  |  |  | X |
| EQ-VAS |  |  |  |  |  | X |

<sup>a</sup> continuous monitoring during the 72h intervention
<sup>b</sup> or hospital discharge if < 5 days
<sup>c</sup> monitored hourly/regularly during the 72h intervention
<sup>d</sup> for women of childbearing potential at baseline only
<sup>e</sup> If pulmonary infection is suspected within 72 hours, a chest X-ray or CT scan should be performed.
<sup>f</sup> If decompressive craniectomy will be carried out within 3 days after randomization, the CT/MRI taken before the decompressive craniectomy will be included in this analysis.
<sup>g</sup> maintained for 72h, with $\leq 4$ short breaks/day $\leq 1$ h each.

### Efficacy End Points

#### Primary outcome

The primary outcome is the distribution of modified Rankin Scale (mRS) scores at 90 days after randomization.

#### Secondary outcomes

The secondary efficacy end points include: (1) The proportion of mRS score of 0 to 1 at 90 days; (2) The proportion of mRS score of 0 to 2 at 90 days; (3) The proportion of mRS score of 0 to 3 at 90 days; (4) The NIHSS score change between baseline and 5 to 7 days or at early discharge; (5) European Quality Visual Analogue Scale (EQ-VAS) score at 90 days.

### Safety End Points

The primary safety end point is all-cause mortality rate at 90 days. Secondary safety end points are the following: (1) Pulmonary infection within 72 hours after randomization; (2) sICH and (3) any ICH, according to modified Heidelberg criteria^17^; (4) cerebral edema(CE) and (5) malignant cerebral edema(MCE) within 72h^18,19^.

### Sample Size Estimates

The sample size estimation for this study is primarily based on the difference in percentage of 90-day mRS of 0 to 2 scores between the head elevation and lying flat group. We assume that the cumulative proportion of favorable functional outcome (defined as an mRS score of 0–2) would be 30.8% in the head elevation group and 23.8% in the control group^20^. The type I error (α) for this trial was set as 0.05 (two-sided), with a power(1-β) of 0.8. The overall sample size was to be 1,332 cases, with a 5% loss-to-follow-up rate taken into account. Therefore, based on a 1:1 randomization ratio, a sample size of 666 patients for each group was determined to detect the pre-defined superiority with a power of 80% and two-sided alpha of 0.05^21^. This calculation was conducted using PASS software (NCSS, LLC. Kaysville, Utah, USA) version 15.0.

### Statistical Analysis

Efficacy outcomes will be analyzed in the intention-to-treat population, with adjustment for age, baseline NIHSS score, baseline ASPECTS, occlusion site, and the time from last known well to randomization. The primary analysis will assess intergroup differences in the distribution of global disability. Primary outcome will be compared using the generalized odds ratio (GenOR). For binary outcomes, the modified Poisson regression with robust error estimation will be employed. Continuous outcomes, such as the EQ-VAS score, will be analyzed by the win ratios^22,23^. Both unadjusted and IPTW adjusted treatment estimates will be reported alongside their corresponding 95% confidence intervals. All statistical tests will be two-sided, with a significance level set at α = 0.05. All analysis will be performed using the SAS version 9.4 (SAS Institute) and R Version 4.3.0 (R Foundation for Statistical Computing). Detailed statistical analyses will be described in the statistical analysis plan which will be finalized before the database lock.

### Data safety monitoring board (DSMB)

In order to guarantee the safety of the patients involved in the study, a DSMB will be established. The DSMB will consist of a neurologist, neuro-interventionalist and statistician, none of whom are involved in the execution of the clinical trial. The DSMB is responsible for recommending to the Steering Committee whether the trial protocol should be revised or the trial should be terminated due to safety concerns.

## DISCUSSION

Head elevation is a simple, low-cost intervention with the potential to influence post-stroke outcomes. Previous studies, including the HeadPoST trial, have explored head elevation in AIS patients but yielded inconclusive results, largely due to the heterogeneity in stroke severity and occlusion type. Furthermore, these trials often excluded EVT-treated patients, who represent a unique subgroup with severe stroke, disrupted blood-brain barrier integrity, and are at high risk of pulmonary infections and CE^24^. While preclinical studies suggest that lying flat increases cerebral blood flow and perfusion in ischemic regions, this benefit has not translated to improved outcomes in clinical trials.

The HeadSOAR trial specifically addresses this evidence gap by focusing on patients with anterior circulation LVO who have undergone EVT with successful reperfusion. This study hypothesizes that head elevation at 30°-40° could reduce complications such as pulmonary infections, ICH, and CE, thereby improving functional outcomes at 90 days. Patients with baseline ASPECTS scores of 8 to 10 were excluded because these patients are expected to do well with lower likelihood of CE and hemorrhagic complications^25^. As more patients with large ischemic core are being treated with EVT^26-28^, this trial does not have a lower limit on the extent of ischemic core for inclusion of patients. Although the trial excludes posterior circulation strokes and lacks intracranial pressure (ICP) monitoring due to logistical and pragmatic constraints, its multicenter design ensures broad applicability. Future research incorporating ICP assessments and exploring head elevation in other stroke subtypes may clarify the role of head elevation in stroke care.

The HeadSOAR trial recruited its initial participant on November 15, 2023. As of January 23, 2025, a total of 1368 subjects have been enrolled in the trial. The study is projected to reach completion, including the collection of 90-day outcomes, by May 2025.

## Summary and Conclusion

In conclusion, the HeadSOAR trial aims to evaluate the impact of head elevation versus a flat lying position on reducing complications and improving neurological outcomes in patients with anterior circulation LVO treated with EVT. This study will provide valuable insights for post-EVT management strategies in clinical practice.

## Data Availability

All data produced in the present study are available upon reasonable request to the authors after 5 years.

